# Public Perceptions of Neurotechnologies Used to Target Mood, Memory, and Motor Symptoms

**DOI:** 10.1101/2024.06.09.24308176

**Authors:** Rémy A. Furrer, Amanda R. Merner, Ian Stevens, Peter Zuk, Theresa Williamson, Francis X. Shen, Gabriel Lázaro-Muñoz

## Abstract

**Background:** Advances in the development of neurotechnologies have the potential to revolutionize treatment of brain-based conditions. However, a critical concern revolves around the willingness of the public to embrace these technologies, especially considering the tumultuous histories of certain neurosurgical interventions. Therefore, examining public attitudes is paramount to uncovering potential barriers to adoption ensuring ethically sound innovation.

**Methods:** In the present study, we investigate public attitudes towards the use of four neurotechnologies (within-subjects conditions): deep brain stimulation (DBS), transcranial magnetic stimulation (TMS), pills, and MRI-guided focused ultrasound (MRgFUS) as potential treatments to a person experiencing either mood, memory, or motor symptoms (between-subjects conditions). US-based participants (N=1052; stratified to be nationally representative based on sex, race, age) were asked about their perceptions of risk, benefit, invasiveness, acceptability, perceived change to the person, and personal interest in using these neurotechnologies for symptom alleviation.

**Results:** Descriptive results indicate variability between technologies that the U.S. public is willing to consider if experiencing severe mood, memory, or motor symptoms. The main effect of neurotechnology revealed DBS was viewed as the most invasive and risky treatment and was perceived to lead to the greatest change to who someone is as a person. DBS was also viewed as least likely to be personally used and least acceptable for use by others. When examining the main effects of symptomatology, we found that all forms of neuromodulation were perceived as significantly more beneficial, acceptable, and likely to be used by participants for motor symptoms, followed by memory symptoms, and lastly mood symptoms. Neuromodulation (averaging across neurotechnologies) was perceived as significantly riskier, more invasive, and leading to a greater change to person for mood versus motor symptoms; however, memory and motor symptoms were perceived similarly with respect to risk, invasiveness, and change to person.

**Conclusion:** These results suggest that the public views neuromodulatory approaches that require surgery (i.e., DBS and MRgFUS) as riskier, more invasive, and less acceptable than those that do not. Further, findings suggest individuals may be more reluctant to alter or treat psychological symptoms with neuromodulation compared to physical symptoms.

## Introduction

Recent decades have seen the emergence of medical neurotechnologies aimed at offering treatments for a wide spectrum of brain-based conditions that affect people’s mood, memory, or motor functions across a variety of methods including ablation, electromagnetic stimulation, and pharmacological neuromodulation.^1^ Given the growing commercial interests in neural implants (e.g., Neuralink), and increasingly promising research on these medical treatments—with the market projections estimating the industry growing to 17 billion annually—it is important to understand the perspectives of the potential end-users (i.e., members of the public).^2–6^ The public’s attitudes may vary drastically given the perceived novelty of these technologies, and may also be influenced by how these technologies are portrayed in the media.^7,8^ Furthermore, given that the early history of some of these neurotechnologies is clouded by ethical controversy and outright abuses of vulnerable patient populations (e.g., the use of subcortical “septal” stimulation in a homosexual patient with comorbid schizophrenia by Dr. Robert Heath as an early form of conversion therapy), it is unclear if there is significant public interest in engaging with these treatments regardless of how effective the treatments have become.^9–13^ It is therefore vital that the next generation of technological innovations for treating brain-based conditions incorporate the views of the public/end-users they aim to help to avoid repeating any past transgressions and ensure ethically sound innovation.

Neurotechnologies vary widely with respect to their mode of treatment delivery; deep brain stimulation (DBS) requires one or more electrodes be implanted into specific brain regions for electrical stimulation, transcranial magnetic stimulation (TMS) induces intracranial effects from the application of a magnetic field on a patient’s scalp, magnetic resonance imaging guided focused ultrasound (MRgFUS) lesions subcortical targets without the use of an open surgical approach (also referred to as “incision-less surgery”), and pharmacological regimens (i.e., pills), require ingesting medications orally to diffusely target an array of mechanisms in the brain.^2,14^ Public perceptions of these technologies may vary given the differences in treatment delivery methods, which may in turn influence how receptive members of the public are to a specific technology.

When it comes to medical neurotechnologies, there has been a particular focus in the academic literature on DBS, which may be in part due to the neuroethical debate around whether, and to what degree, DBS may impact a patient’s personality and related characteristics.^15–21^ There have been numerous evaluations of psychosocial impacts of DBS devices for psychiatric and movement disorders over the past 10-15 years.^16–49^ There have also been some qualitative studies of TMS treatments during this time;^50–53^ however, only recently has there been a focused examination of MRgFUS.^54^ Previous investigations of neurotechnologies have tended to fall into silos with respect to the disorders or symptomatology being studied, namely those researchers focused on motor (i.e., “doing”) symptoms^23,27,31,33,34,43,45,49,55–58^ or those involving mood/psychiatric (i.e., “feeling”), ^14,26,40,46,52,54,59–63^ or memory/cognition (i.e., “thinking”).^35,64–66^ However, research simultaneously examining varying forms of neuromodulation as well as how the type of disorder being treated may influence views on the technologies is lacking. Finally, much of the previous work has been limited to examining the perspectives of patients and clinicians, and the studies that did explore the public’s attitudes around some of these neuromodulatory interventions were conducted using either surveys or assessments of social media.^29,41,53,59,67–69^

We aim to address several of the meaningful gaps in the literature identified above. First, our study employs an experimental approach to comparatively examine the public attitudes towards neurotechnologies, which expands on previous work that was limited to observational work, (i.e., surveys or media analyses). We also include a range of both neurotechnologies (i.e., DBS, MRgFUS, TMS, and pills) and symptomatology (i.e., mood, memory, and motor) to address the need for work that spans the siloed research on this topic. Findings from this study provide critical insight into the complex public attitudes related to risks, benefits, invasiveness, acceptability, perceived changes to the patient, and the likelihood of personal use across a range of neurotechnologies. These findings can help identify barriers to uptake of these technologies, aid clinicians in addressing public perceptions surrounding these technologies, and inform the responsible development and use of these and future neurotechnologies.

## Methods

### Overview

We used an experimental design to examine public attitudes towards the use of four neurotechnologies: DBS, TMS, Pills, and MRgFUS. These technologies were presented to participants as potential treatments being offered to a person described as experiencing symptoms severely affecting one of three brain functions: mood, memory, or motor.

### Participants

Participants were recruited from Prolific, an online sampling firm, using the platform’s nationally representative stratified sampling option using self-reported age, gender, and race demographics. All participants provided informed consent prior to participating in the experiment. All research activities were approved by the Harvard Medical School Institutional Review Board [IRB22-0986].

### Procedure

All participants rated each neurotechnology (4 within-subjects conditions: DBS, TMS, Pills, MRgFUS) based on a target person experiencing one of three symptoms (3 between-subjects conditions: Mood, Memory, Motor).

### Description of Symptoms

Participants were presented with the following descriptions of the target’s **symptoms**: “A person has been experiencing the following:”

**Mood symptoms** (e.g., feeling sad, irritable, empty), a loss of pleasure or interest in activities, for most of the day, every day. They experience poor concentration, feelings of excessive low self-worth, hopelessness about the future, disrupted sleep, changes in appetite, and feeling tired.

**Memory symptoms** (e.g., unable to recall memories, difficulty retaining new information), memory loss for most of the day, every day. They experience difficulty learning and recalling new information such as recent events, conversations, or people, and recalling important memories and personal information about themselves.

**Motor symptoms** (e.g., slowed movement, muscle weakness), a loss of muscle control, for most of the day, every day. They experience tremors while their muscles are at rest, stiffness, trouble swallowing, unstable posture, difficulties with walking, and reduced control over their facial muscles.

### Description of Neurotechnologies

Following the description of the target person’s symptoms, participants were presented with the following descriptions of the neurotechnologies: “Given the severity of their condition, they are presented with the following **neurotechnology** to help reduce symptoms:”

**Deep Brain Stimulation** (DBS) which involves surgically implanting electrodes into the brain to deliver electrical stimulation to a specific region of the brain.

**Transcranial Magnetic Stimulation** (TMS) which involves placing a magnet against an area (outside) of the head to deliver magnetic stimulation to a specific region of the brain.

**Pills** which involve ingesting medication (taken by mouth) in the form of a pill to deliver chemicals to the brain.

**MRI-guided focused ultrasound** (MRgFUS) which involves placing a cap on the outside of the head that delivers focused sound waves to create a precise lesion in a specific region of the brain.

### Dependent measures

Following the between-subjects assignment to one of the three symptom conditions (Mood, Memory, or Motor symptoms), all participants were asked to respond to questions about the six dependent variables for each of the four (within-subjects) neurotechnologies: “Given this person’s (Mood / Memory / Motor) symptoms, to what extent do you think using (DBS / TMS / Pills / MRgFUS) would be (beneficial / risky / invasive / acceptable)?” on 5-point Likert Scales (range: *1= Not at all, 5= Extremely*). Upon answering the four dependent variables mentioned above, participants answered two more questions for each of the neurotechnologies: “Given this person’s (Mood / Memory / Motor) symptoms, to what extent do you think using (DBS / TMS / Pills / MRgFUS) would change who they are as a person?” on 5-point Likert Scales (*1 = Not at all, 5= A great deal*) and finally, “Now, suppose YOU were experiencing these (Mood / Memory / Motor) symptoms, would you consider (DBS / TMS / Pills / MRgFUS)?” on 5-point Likert Scales (range: *1= I definitely would not, 5= I definitely would*). The above-mentioned measures represent an initial subset of questions presented to participants. Additional questions about these Neurotechnologies were asked at the end of the survey for a separate manuscript.

### Analyses

We ran a mixed-effects ANOVA on each of the six dependent variables: Benefit, Acceptability, Personal Use, Risk, Invasiveness, and Change to Person. Mauchly’s test of sphericity indicated that the assumption of sphericity was violated (*p* < .001) for each outcome variable, therefore we applied Huynh-Feldt sphericity corrections for each analysis. Significant main effects were followed up with Bonferroni-corrected post hoc comparisons.

## Results

### Participants

Of the 1,145 participants who began the study by signing the consent form, 61 participants were removed because they failed either the initial bot check or the attention check. Of the remaining participants, 1,052 completed all the main dependent measures. Participants were almost evenly split with respect to gender (n females = 514; n males = 507), were predominantly White (75%), with an average age of 45.5 years (see Table 1 for detailed demographic information).

**Figure 1.**
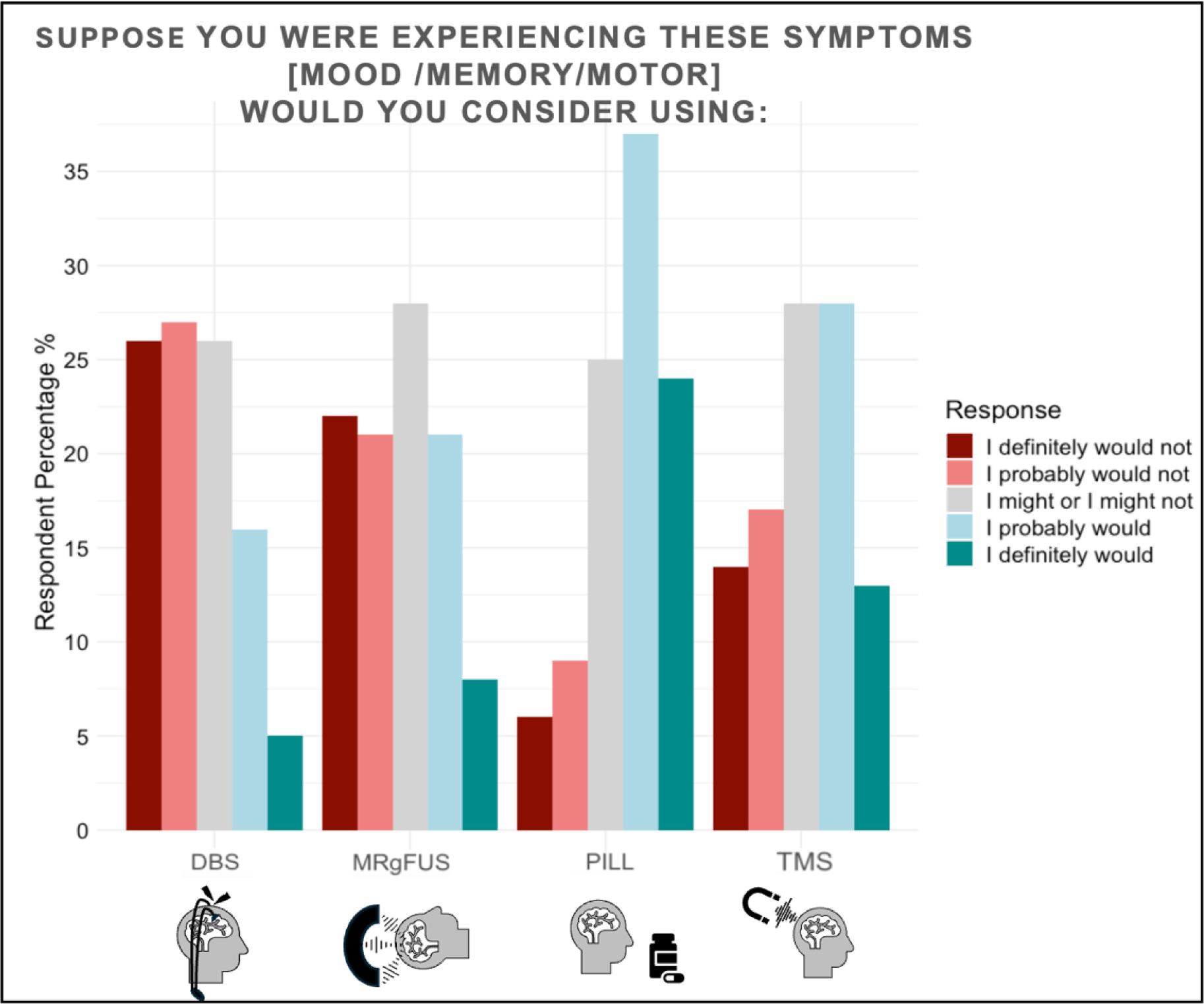
Descriptive results of participant likelihood of using neurotechnologies (averaged over symptoms) ***Note:*** Participants rated their likelihood of using each of the four neurotechnologies on the x-axis on a 5-point Likert scale. The y-axis represents the percentage of participants who selected each answer for each neurotechnology. These results represent participants’ reported likelihood of using each technology averaged over the three potential symptoms (mood, memory, and motor). Significant variation in reported likelihood to use these neurotechnologies is observed based on symptoms, which is further examined in Figure 3.

**Table 1.** Sample Demographics.

| **Table 1. Sample Demographics** |  |  |
| --- | --- | --- |
| **Sample size (N)** |  | 1084 |
| **Age** |  |  |
| Mean (SD) |  | 45.5 (SD=16.1) |
| **Gender** |  |  |
| Female |  | 514 (49.1%) |
| Male |  | 507 (48.5%) |
| Trans female/Trans woman |  | 3 (0.28) |
| Trans male/Trans man |  | 7 (0.7%) |
| Genderqueer/Gender non-conforming |  | 12 (1.1%) |
| *Other |  | 25 (2.4%) |
| **Race** |  |  |
| American Indian, Native American, Alaska Native |  | 11 (1%) |
| Asian |  | 65 (6%) |
| Black or African American |  | 135 (12.5%) |
| Native Hawaiian, Pacific Islander |  | 3 (0.3%) |
| †Other |  | 8 (0.7%) |
| White |  | 825 (76.1%) |
| **Ethnicity** |  |  |
| Not Hispanic or Latino |  | 992 (91.5%) |
| Hispanic or Latino |  | 57 (5.3%) |
| **Education level** |  |  |
| Bachelor's or higher |  | 551 (52.6%) |
| **Household income** |  |  |
| \$0–\$49,999 | | 444 (42.4%) |
| ≥ \$50,000–109,999 | | 399 (36.8%) |
| ≥ \$110,000 | | 205 (18.9%) |

\*Other gender in sample 1 included the following self-reported identities: Non-Binary, trans nonbinary genderfluid, transmasculine. †Other race/ethnicity included the following self-reported races (counts): Middle Eastern (2), Mediterranean (1), Jewish (2), Indigenous American (1), Hebrew (1) French / Indian / Black / White (1). Note: Participants could select multiple races, therefore, percentages sum above 100%. Missing values across demographics ranged from 35-38.

#### Benefit, Acceptability, and Personal Use

Analyses revealed a significant main effect of Neurotechnology on Benefit (*F*(2.92, 1049) = 83.24, *p* < .001, ηp² = .07), Acceptability (*F*(2.88, 1049) = 318.53, *p* < .001, ηp² = .23), and Personal Use (*F*(2.90, 1049) = 336.53, *p* < .001, ηp² = .24). See Figure 2 for plots. Post hoc tests revealed that Pills were rated as most beneficial (Mean= 3.33, SD= 0.92), followed by DBS (Mean= 3.03, SD= 0.96), and then TMS (Mean= 2.88, SD= 1.01) and MRgFUS (Mean= 2.88, SD= 1.06). All post hoc comparisons for perceived Benefit were significant (*p* < .001) except for TMS and MRgFUS (*p* = 1.00). Post hoc tests revealed that, for Acceptability and Personal Use, Pills were rated as most acceptable (Mean= 3.75, SD= 1.01) and most likely to be used by participants (Mean= 3.65, SD= 1.11), followed by TMS (Acceptability: Mean= 3.22, SD= 1.07; Personal Use: Mean= 3.08, SD= 1.23), then MRgFUS (Acceptability: Mean= 2.84, SD= 1.13; Personal Use: Mean= 2.71, SD= 1.24), and finally DBS (Acceptability: Mean= 2.72, SD= 1.03; Personal Use: Mean= 2.47, SD= 1.18). All post hoc comparisons were significant (*p* < .005).

**Figure 2.**
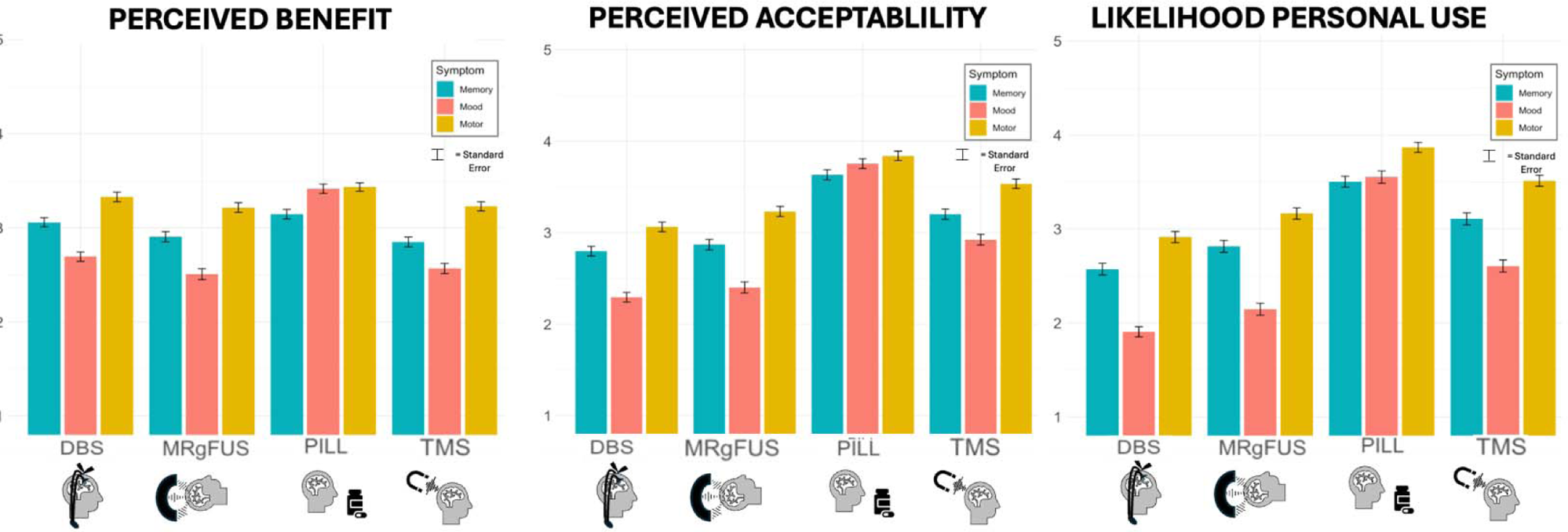
Main Effect of Neurotechnology.

#### Risk, Invasiveness, and Change to Person

The results of the ANOVAs revealed a significant main effect of Neurotechnology on Risk (*F*(2.91, 1049) = 570.80, *p* < .001, ηp² = .35), Invasiveness (*F*(2.92, 1049) = 1027.31, *p* < .001, ηp² = .50), and Change to Person (*F*(2.93, 1049) = 164.62, *p* < .001, ηp² = .14). See Figure 3 for plots. Post hoc tests revealed that for Risk and Change to Person, participants perceived DBS as the riskiest (Mean= 3.83, SD= 1.04) and leading to the greatest change to the person (Mean= 2.69, SD= 1.13), followed by MRgFUS (Risk: Mean= 3.39, SD= 1.19; Change to Person: Mean= 2.52, SD= 1.12), Pills (Risk: Mean= 2.54, SD= 0.89; Change to Person: Mean= 2.26, SD= 1.04), and TMS (Risk: Mean= 2.40, SD= 1.09; Change to Person: Mean= 2.02, SD= 1.04). All post hoc comparisons were significant (*p* < .002). Post hoc tests for Invasiveness revealed that participants perceived DBS as the most invasive (Mean= 4.16, SD= 1.04), followed by MRgFUS (Mean= 3.18, SD= 1.31), TMS (Mean= 2.13, SD= 1.15), and lastly Pills (Mean= 1.94, SD= 1.00). All post hoc comparisons between the four Neurotechnologies were significant (*p* < .001).

**Figure 3.**
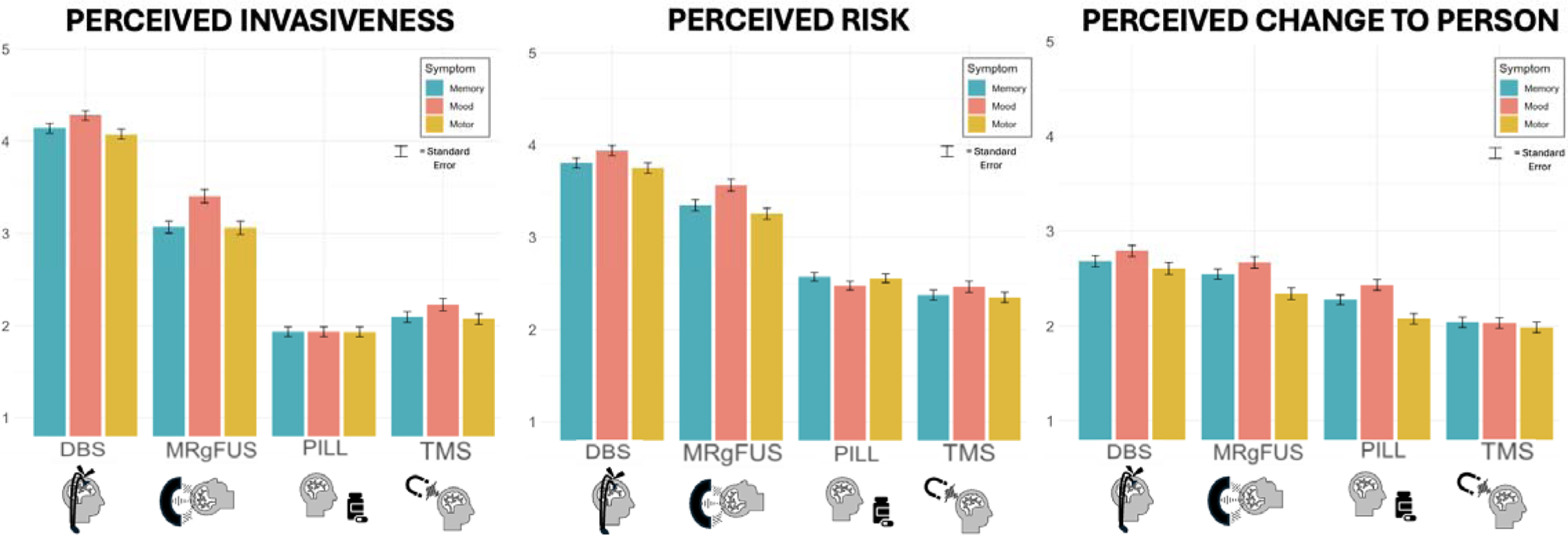

### Main Effect of Symptoms

#### Benefit, Acceptability, and Personal Use

The results of the ANOVAs revealed a significant main effect of Symptoms on Benefit (*F*(2, 1049) = 47.34, *p* < .001, ηp² = .08), Acceptability (*F*(2, 1049) = 55.54, *p* < .001, ηp² = .10), and Personal Use (*F*(2, 1049) = 86.09, *p* < .001, ηp² = .14). Post hoc tests revealed that the average effect of Neurotechnology was rated as most beneficial for Motor symptoms (Mean= 3.30, SD= 0.93), followed by Memory symptoms (Mean= 2.99, SD= 0.96), and finally Mood symptoms (Mean= 2.80, SD= 1.05). Similarly, Neurotechnology was rated as most acceptable for Motor symptoms (Mean= 3.42, SD= 1.05), followed by Memory symptoms (Mean= 3.13, SD= 1.08), and Mood symptoms (Mean= 2.85, SD= 1.19). For Personal Use, Neurotechnology was also rated as most likely to be used for Motor symptoms (Mean= 3.37, SD= 1.16), followed by Memory symptoms (Mean= 3.00, SD= 1.20), and Mood symptoms (Mean= 2.56, SD= 1.32). All post hoc comparisons between the three symptoms were significant (*p* < .001).

#### Risk, Invasiveness, and Change to Person

The results of the ANOVAs revealed a significant main effect of Symptoms on Risk (*F*(2, 1049) = 3.53, *p* = .03, ηp² = .01), Invasiveness (*F*(2, 1049) = 7.04, *p* < .001, ηp² = .01), and Change to Person (*F*(2, 1049) = 6.31, *p* = .002, ηp² = .01). Post hoc tests revealed that the average effect of Neurotechnology was rated as significantly riskier (*p* = .03, Mean= 3.11, SD= 1.27), more invasive (*p* < .002, Mean= 2.96, SD= 1.50), and leading to a greater change to person (*p* = .001, Mean= 2.48, SD= 1.13) for Mood symptoms compared to Motor symptoms (Risk: Mean= 2.98, SD= 1.19; Invasiveness: Mean= 2.78, SD= 1.43; Change to Person: Mean= 2.25, SD= 1.15). Post hoc differences between Mood and Memory symptoms were significant for Invasiveness (*p* = .009, Mood: Mean= 2.96, SD= 1.50; Memory: Mean= 2.81, SD= 1.39), but not for Risk (*p* = 0.27, Mood: Mean= 3.11, SD= 1.27; Memory: Mean= 3.02, SD= 1.18), or Change to Person (*p* = .45, Mood: Mean= 2.48, SD= 1.13; Memory: Mean= 2.39, SD= 1.04). There were no significant post hoc differences between Motor and Memory symptoms for Risk (*p* = 1.00), Invasiveness (*p* = 1.00), or Change to Person (*p* = .11).

### Interaction effects of Neurotechnology and Symptom

The results of the ANOVAs showed significant interaction effects for Benefit (*F*(5.85, 1049) = 20.92, *p* < .001, ηp² = .04), Acceptability (*F*(5.76, 1049) = 16.82, *p* = .001, ηp² = .03), Personal Use (*F*(5.80, 1049) = 16.30, *p* = .001, ηp² = .03), Risk (*F*(5.82, 1049) = 3.11, *p* = .005, ηp² = .01), Invasiveness (*F*(5.84, 1049) = 2.12, *p* = .05, ηp² = .004), and Change to Person (*F*(5.86, 1049) = 3.42, *p* = .003, ηp² = .01). To further investigate the interaction effects, we conducted a series of post hoc tests with Bonferroni corrections (correcting for 66 total estimates). Due to space limitations, we report the results that speak to the primary findings in the manuscript, and the full list of results for each post hoc test can be found in supplemental materials.

#### Benefit, Acceptability, and Personal Use

As depicted in Figure 2, perceived benefit results for Symptoms across technologies were similar except for Pills being perceived as significantly more beneficial for Mood compared to Memory symptoms *p* < .001, but not significantly different for Motor symptoms *p* = 1.00. Pills were perceived as significantly more beneficial in treating Mood symptoms compared to any of the other three neurotechnologies. The pattern of results for symptoms across technologies were similar except for Pills, for which there were no significant differences across symptoms (*p* = 1.00), because Pills were perceived as particularly more acceptable to treat Mood symptoms compared to the other Neurotechnologies. As depicted in Figure 2, the pattern of results for Symptoms across technologies were similar except for Pills being perceived as significantly more likely to be used for Motor symptoms compared to Mood (*p* = .010) and Memory (*p* < .001). Pills were rated as being significantly more likely to be personally used to treat mood symptoms compared to the other neurotechnologies.

#### Risk, Invasiveness, Change to Person

As depicted in Figure 3, the pattern of risk perceptions for symptoms across technologies were similar, with Mood > Memory > Motor except for Pills where there were no significant differences across symptoms (*p* = 1.00). The pattern of results for perceived Invasiveness for Symptoms across technologies were similar to Risk perceptions, except for Pills where there were no significant differences across symptoms (*p* = 1.00). Perceptions of Change to Person for Symptoms across technologies were similar for Risk and Invasiveness, except that there were no significant differences across symptoms for DBS and TMS (*p* = 1.00), but Motor compared to Mood symptoms were perceived as resulting in less change to the person for Pills (*p* < .001) and MRgFUS (*p* = .006).

## Discussion

This study examined participants’ perceptions of several forms of neuromodulation across three symptom profiles (i.e., mood, memory, motor). Our findings contribute to the handful of prior analyses of public opinion surrounding the use of neurotechnologies to treat brain-based conditions.^29,41,53,59,67,69,70^ Descriptive results indicate variability around the technologies that the U.S. public is willing to consider if experiencing severe mood, motor, or memory symptoms. Specifically, 21% of respondents would “probably” use or “definitely” use DBS (26% uncertain), 29% for MRgFUS (28% uncertain), 41% for TMS (28% uncertain), and 61% for pills (25% uncertain). These findings suggest potential openness to adopting neurotechnological interventions among the general population. Although there were broad patterns that emerged (e.g., DBS and MRgFUS were seen as more invasive, risky, and causing more change to person than Pills or TMS), there were notable nuances in participants’ views on these neurotechnologies and their utility for different types of conditions.

DBS is an established intervention for treatment of severe movement disorders and has shown promise for some treatment-refractory psychiatric conditions.^71^ However, despite DBS being perceived as more beneficial than MRgFUS and TMS, it was also rated as less acceptable and less likely to be used. DBS was also viewed by participants as the riskiest and most invasive neurotechnology. This suggests that despite its efficacy and potential benefit, the public still has concerns about this treatment approach. This is consistent with some previous work showing the public holds generally positive, but cautious, views of DBS.^29,59,69^

When considered alongside MRgFUS, which produces a permanent lesion, DBS offers greater flexibility as a treatment option—with titratable stimulation parameters and the ability to be removed in the event of complications or lack of efficacy.^72^ Therefore, from a clinical perspective, it may be surprising that participants rated DBS as more invasive and riskier than MRgFUS. However, participants’ responses may be influenced by the implantation process, as DBS requires an “open surgical approach” for placement of the device, as well as the ongoing presence of the device in the individual’s body.

Participants also viewed DBS as being significantly more likely to change someone as a person than the other technologies, which aligns with the ongoing debate within the neuroethics literature.^15,16,18–20,30,46,47^ Concerns have been raised that media coverage of this debate may contribute to the public’s views of acceptability toward DBS, specifically.^73^

Participants’ views on pills stand in contrast to those of DBS, with this form of neuromodulation being rated as the most beneficial, acceptable, and likely to be used. This trend may be, at least in part, related to how common oral medications are in the U.S., particularly those used to modulate mood (e.g., antidepressants, anxiolytics).^74–76^ Pills were also perceived as the least invasive neurotechnology, despite qualitative findings noting the potential for medications to be perceived as invasive given their systemic effects on the body.^77^ It is notable that while pills were viewed as the least invasive, they were also viewed as riskier and as leading to greater change to person than TMS. Yet participants still felt that pills were more acceptable and were more open to using them than TMS. This aligns with previous work demonstrating that people hold generally positive views of TMS but may only be willing to consider it as a treatment option if medication are not effective.^59^

Across all four neurotechnologies, participants felt these treatments were most beneficial, acceptable, and were more likely to use them personally in the context of motor symptoms, followed by memory and mood symptoms. This is consistent with a previous media analysis examining DBS, which found that the use of DBS for movement disorders was viewed as a more effective treatment for movement disorders than psychiatric disorders (64% versus 9%).^41^ We found that the use of these technologies to treat mood symptoms was viewed to carry the greatest levels of risk, invasiveness, and change to person. This distinction between acceptability of neuromodulation for brain-based conditions that manifest through physical symptoms versus psychological symptoms will be critical to understand in greater detail, including the range of reasons for which respondents hold these judgments. The reasons underlying these judgments may have important implications for potential adoption of these technologies in psychiatry and reduction of stigma for individuals who are already engaging with these interventions. Finally, the pattern of results on perceived acceptability most closely resembled the pattern of results for personal use, which suggests that perceived norms may play a role in the potential uptake of these technologies.

## Limitations

To examine the potential impact of different symptomatology on the public’s views of these neurotechnologies, we were limited to selecting technologies for which there are applications across all of the symptom domains (i.e., mood, motor, memory). Based on previous work, we acknowledge that there are some neuromodulation approaches (e.g., electroconvulsive therapy (ECT)) that may have elicited different responses given the stigma and long history of this technology in psychiatry and recognize the absence of this technology from the experiment is a notable limitation.^78^ In addition, we did not examine neuromodulation via digital tools, nor strategies that do not rely on technology, such as cognitive behavioral therapy. In the current study, we also opted for respondents to subjectively interpret the outcomes of interest (e.g., invasiveness, change to person). Given a lack of consensus on the definition of these constructs and the different types of invasiveness observed in the context of neuromodulation, we opted not to provide specific definitions.^77^ Future projects interested in additional nuances within these constructs could use these results as an intuitive baseline and examine how providing specific definitions might influence opinions. Finally, we note that participants were questioned about their interest in using these technologies in the absence of any information about the costs associated with each intervention. Given the substantial financial costs associated with many of these treatments, this is an important potential barrier to access and uptake that should be considered in future work.^79–81^

## Conclusion

As rates of brain-based conditions continue to rise, members of the public may one day stand to benefit from some form of neuromodulation included in the present study. Our results suggest that despite viewing interventions as effective and potentially beneficial, the public views some forms of neuromodulation as invasive, risky, and able to change who they are as a person, which may deter individuals from engaging with them. These findings provide novel insights into the public’s complex views on neuromodulation, which can be used to facilitate conversations about barriers to uptake, ethical safeguards on novel applications of neuromodulation, and alignment of technology development and use with the values of end-users. This engagement will serve, we hope, to maximize the available benefits of current and future neurotechnologies while minimizing the risks through careful collective deliberation.

## Data Availability

All data produced in the present study are available upon reasonable request to the authors

## Funding

This research was funded in part by the DANA Foundation. PZ’s contribution to the research reported in this publication was also supported by the National Institute of Neurological Disorders and Stroke (NINDS) of the National Institutes of Health (NIH) as part of the NIH BRAIN Initiative under Award Number F32MH127776, administered by the National Institute of Mental Health (NIMH). The content is solely the responsibility of the authors and does not necessarily represent the official views of the NIH, BRAIN Initiative, NINDS, NIMH, Harvard Medical School, Minnesota Law School, or MGH

## Disclosures

Authors have no conflicts of interest or disclosures to report.

## Notes

### Competing Interest Statement

The authors have declared no competing interest.

### Funding Statement

This research was funded in part by the DANA Foundation. PZ contribution to the research reported in this publication was also supported by the National Institute of Neurological Disorders and Stroke (NINDS) of the National Institutes of Health (NIH) as part of the NIH BRAIN Initiative under Award Number F32MH127776, administered by the National Institute of Mental Health (NIMH). The content is solely the responsibility of the authors and does not necessarily represent the official views of the NIH BRAIN Initiative, NINDS, NIMH, Harvard Medical School, Minnesota Law School, or MGH

### Author Declarations

Approved by Harvard Medical School Institutional Review Board

## References

1. Denison, T. & Morrell, M. J. Neuromodulation in 2035. Neurology 98, 65–72 (2022).

2. Pomeraniec, J., Elias, W. J. & Moosa, S. High-Frequency Ultrasound Ablation in Neurosurgery. Neurosurg. Clin. N. Am. 34, 301–310 (2023).

3. Neurotech Reports. https://www.neurotechreports.com/pages/execsum.html.

4. Arulchelvan, E. & Vanneste, S. Promising neurostimulation routes for targeting the hippocampus to improve episodic memory: A review. Brain Res. 1815, 148457 (2023).

5. Xu, G., Li, G., Yang, Q., Li, C. & Liu, C. Explore the durability of repetitive transcranial magnetic stimulation in treating post-traumatic stress disorder: An updated systematic review and meta-analysis. Stress Health J. Int. Soc. Investig. Stress (2023) doi:10.1002/smi.3292.

6. Matsugi, A., Ohtsuka, H., Bando, K., Kondo, Y. & Kikuchi, Y. Effects of non-invasive brain stimulation for degenerative cerebellar ataxia: a protocol for a systematic review and meta-analysis. BMJ Open 13, e073526 (2023).

7. Purcell-Davis, A. The Representations of Novel Neurotechnologies in Social Media. New Bioeth. (2015) doi:10.1179/2050287713Z.00000000026.

8. Racine, E., Waldman, S., Rosenberg, J. & Illes, J. Contemporary neuroscience in the media. Soc. Sci. Med. 71, 725–733 (2010).

9. Hariz, M. I., Blomstedt, P. & Zrinzo, L. Deep brain stimulation between 1947 and 1987: the untold story. Neurosurg. Focus 29, E1 (2010).

10. Spiegel, E. A., Wycis, H. T., Marks, M. & Lee, A. J. Stereotaxic Apparatus for Operations on the Human Brain. Science 106, 349–350 (1947).

11. Baumeister, A. A. The Tulane Electrical Brain Stimulation Program a historical case study in medical ethics. J. Hist. Neurosci. 9, 262–278 (2000).

12. Fins, J. J. From psychosurgery to neuromodulation and palliation: history’s lessons for the ethical conduct and regulation of neuropsychiatric research. Neurosurg. Clin. N. Am. 14, 303–319 (2003).

13. Nadler, R. & Chandler, J. A. Legal Regulation of Psychosurgery: A Fifty-State Survey. J. Leg. Med. 39, 335–399 (2019).

14. Steuber, E. R. & McGuire, J. F. A Meta-Analysis of Transcranial Magnetic Stimulation in Obsessive Compulsive Disorder. Biol. Psychiatry Cogn. Neurosci. Neuroimaging S2451-9022(23)00146–5 (2023) doi:10.1016/j.bpsc.2023.06.003.

15. Gilbert, F., M. Viana, J. N. & Ineichen, C. Deflating the Deep Brain Stimulation Causes Personality Changes Bubble: the Authors Reply. Neuroethics 14, 125–136 (2021).

16. Thomson, C. J., Segrave, R. A. & Carter, A. Changes in Personality Associated with Deep Brain Stimulation: a Qualitative Evaluation of Clinician Perspectives. Neuroethics 14, 109–124 (2021).

17. Wilt, J. A., Merner, A. R., Zeigler, J., Montpetite, M. & Kubu, C. S. Does Personality Change Follow Deep Brain Stimulation in Parkinson’s Disease Patients? Front. Psychol. 12, (2021).

18. Merner, A. R. et al. Participant perceptions of changes in psychosocial domains following participation in an adaptive deep brain stimulation trial. Brain Stimulat. 16, 990–998 (2023).

19. Kubu, C. S. et al. Pragmatism and the Importance of Interdisciplinary Teams in Investigating Personality Changes Following DBS. Neuroethics 14, 95–105 (2021).

20. Merner, A. R., et al. A Patient-Centered Perspective on Personality Change Following Deep Brain Stimulation. SSRN Scholarly Paper at 10.2139/ssrn.4665830 (2023).

21. Zuk, P. et al. Researcher Views on Changes in Personality, Mood, and Behavior in Next-Generation Deep Brain Stimulation. AJOB Neurosci. 14, 287–299 (2023).

22. Apantaku, G. O. et al. Clinician preferences for neurotechnologies in pediatric drug-resistant epilepsy: A discrete choice experiment. Epilepsia 63, 2338–2349 (2022).

23. Austin, A., Lin, J.-P., Selway, R., Ashkan, K. & Owen, T. What parents think and feel about deep brain stimulation in paediatric secondary dystonia including cerebral palsy: A qualitative study of parental decision-making. Eur. J. Paediatr. Neurol. 21, 185–192 (2017).

24. Bell, E., Maxwell, B., McAndrews, M. P., Sadikot, A. & Racine, E. Hope and Patients’ Expectations in Deep Brain Stimulation: Healthcare Providers’ Perspectives and Approaches. J. Clin. Ethics (2010) doi:10.1086/JCE201021204.

25. Bell, E., Maxwell, B., McAndrews, M. P., Sadikot, A. & Racine, E. Deep Brain Stimulation and Ethics: Perspectives from a Multisite Qualitative Study of Canadian Neurosurgical Centers. World Neurosurg. 76, 537–547 (2011).

26. Bell, E. & Racine, E. Clinical and ethical dimensions of an innovative approach for treating mental illness: a qualitative study of health care trainee perspectives on deep brain stimulation. Can. J. Neurosci. Nurs. 35, 23–32 (2013).

27. Cabrera, L. Y., Kelly-Blake, K. & Sidiropoulos, C. Perspectives on Deep Brain Stimulation and Its Earlier Use for Parkinson’s Disease: A Qualitative Study of US Patients. Brain Sci. 10, 34 (2020).

28. Cabrera, L. Y., Young Han, C., Ostendorf, T., Jimenez-Shahed, J. & Sarva, H. Neurologists’ Attitudes Toward Use and Timing of Deep Brain Stimulation. Neurol. Clin. Pract. 11, 506–516 (2021).

29. Elkaim, L. M. et al. Deep brain stimulation in children and youth: perspectives of patients and caregivers gleaned through Twitter. Neurosurg. Focus 53, E11 (2022).

30. Gilbert, F., Goddard, E., Viaña, J. N. M., Carter, A. & Horne, M. I Miss Being Me: Phenomenological Effects of Deep Brain Stimulation. AJOB Neurosci. 8, 96–109 (2017).

31. Haahr, A., Kirkevold, M., Hall, E. O. C. & Østergaard, K. ‘Being in it together’: living with a partner receiving deep brain stimulation for advanced Parkinson’s disease – a hermeneutic phenomenological study. J. Adv. Nurs. 69, 338–347 (2013).

32. Haan, S. de, Rietveld, E., Stokhof, M. & Denys, D. Effects of Deep Brain Stimulation on the Lived Experience of Obsessive-Compulsive Disorder Patients: In-Depth Interviews with 18 Patients. PLOS ONE 10, e0135524 (2015).

33. Hariz, G.-M., Limousin, P., Tisch, S., Jahanshahi, M. & Fjellman-Wiklund, A. Patients’ perceptions of life shift after deep brain stimulation for primary dystonia—A qualitative study. Mov. Disord. 26, 2101–2106 (2011).

34. İbrahimoğlu, Ö., Mersin, S. & Akyol, E. The Experiences of Patients with Deep Brain Stimulation in Parkinson’s Disease: Challenges, Expectations, and Accomplishments. Acta Medica Acad. 49, 36–43 (2020).

35. Klein, E. et al. Views of stakeholders at risk for dementia about deep brain stimulation for cognition. Brain Stimulat. 16, 742–747 (2023).

36. Leykin, Y. et al. Participants’ Perceptions of Deep Brain Stimulation Research for Treatment-Resistant Depression: Risks, Benefits, and Therapeutic Misconception. AJOB Prim. Res. 2, 33–41 (2011).

37. Merner, A. R. et al. Changes in Patients’ Desired Control of Their Deep Brain Stimulation and Subjective Global Control Over the Course of Deep Brain Stimulation. Front. Hum. Neurosci. 15, (2021).

38. Mosley, P. E. et al. ‘Woe Betides Anybody Who Tries to Turn me Down.’ A Qualitative Analysis of Neuropsychiatric Symptoms Following Subthalamic Deep Brain Stimulation for Parkinson’s Disease. Neuroethics 14, 47–63 (2021).

39. Brodsky, M. A. et al. Clinical outcomes of asleep vs awake deep brain stimulation for Parkinson disease. Neurology 89, 1944–1950 (2017).

40. Naesström, M., Blomstedt, P., Hariz, M. & Bodlund, O. Deep brain stimulation for obsessive-compulsive disorder: Knowledge and concerns among psychiatrists, psychotherapists and patients. Surg. Neurol. Int. 8, 298 (2017).

41. Robillard, J. M., Cabral, E. & Feng, T. L. Online Health Information-Seeking: The Case of Deep Brain Stimulation in Social Media. Care Wkly. (2018) doi:10.14283/cw.2018.8.

42. Saway, B. F. et al. Medical Students’ Knowledge and Perception of Deep Brain Stimulation. J. Med. Educ. Curric. Dev. 8, 2382120521989977 (2021).

43. Scaratti, C. et al. Long term perceptions of illness and self after Deep Brain Stimulation in pediatric dystonia: A narrative research. Eur. J. Paediatr. Neurol. 26, 61–67 (2020).

44. Stoehr, K. et al. Deep Brain Stimulation in Early-Stage Parkinson’s Disease: Patient Experience after 11 Years. Brain Sci. 12, 766 (2022).

45. Testini, P., Sarva, H., Schwalb, J., Barkan, S. & Cabrera, L. Y. Neurosurgeons perspective on the shift towards earlier use of deep brain stimulation for Parkinson disease. Interdiscip. Neurosurg. 25, 101224 (2021).

46. Thomson, C. J. et al. Personal and relational changes following deep brain stimulation for treatment-resistant depression: A prospective qualitative study with patients and caregivers. PLOS ONE 18, e0284160 (2023).

47. Thomson, C. J. et al. ‘He’s Back so I’m Not Alone’: The Impact of Deep Brain Stimulation on Personality, Self, and Relationships in Parkinson’s Disease. Qual. Health Res. 30, 2217–2233 (2020).

48. Versalovic, E. et al. Deep Brain Stimulation for Substance Use Disorders? An Exploratory Qualitative Study of Perspectives of People Currently in Treatment. J. Addict. Med. (2023) doi:10.1097/adm.0000000000001150.

49. Zhang, C. et al. Deep Brain Stimulation for Parkinson’s Disease During the COVID-19 Pandemic: Patient Perspective. Front. Hum. Neurosci. 15, (2021).

50. Wallman, E. J. et al. Acceptability, safety and tolerability of antidepressant repetitive transcranial magnetic stimulation for adolescents: A mixed-methods investigation. J. Affect. Disord. 310, 43–51 (2022).

51. van Lieshout, E. C., Jacobs, L. D., Pelsma, M., Dijkhuizen, R. M. & Visser-Meily, J. M. Exploring the experiences of stroke patients treated with transcranial magnetic stimulation for upper limb recovery: a qualitative study. BMC Neurol. 20, 365 (2020).

52. Taşdemir Yiğitoğlu, G., Çunkuş Köktaş, N. & Özgün Öztürk, F. Opinions of Depression Patients About Transcranial Magnetic Stimulation: A Qualitative Study. J. Radiol. Nurs. 42, 114–120 (2023).

53. Cabrera, L. Y. & Reiner, P. B. Understanding public (mis)understanding of tDCS for enhancement. Front. Integr. Neurosci. 9, (2015).

54. Asher, R., Hyun, I., Head, M., Cosgrove, G. R. & Silbersweig, D. Neuroethical implications of focused ultrasound for neuropsychiatric illness. Brain Stimulat. 16, 806–814 (2023).

55. Bell, E., Maxwell, B., McAndrews, M. P., Sadikot, A. F. & Racine, E. A Review of Social and Relational Aspects of Deep Brain Stimulation in Parkinson’s Disease Informed by Healthcare Provider Experiences. Park. Dis. 2011, e871874 (2011).

56. Haahr, A. et al. Sharing our story individualized and triadic nurse meetings support couples adjustment to living with deep brain stimulation for Parkinson’s disease. Int. J. Qual. Stud. Health Well-Being 15, 1748361 (2020).

57. Mulroy, E., Robertson, N., Macdonald, L., Bok, A. & Simpson, M. Patients’ Perioperative Experience of Awake Deep-Brain Stimulation for Parkinson Disease. World Neurosurg. 105, 526–528 (2017).

58. Shahmoon, S., Limousin, P. & Jahanshahi, M. Exploring the Caregiver Role after Deep Brain Stimulation Surgery for Parkinson’s Disease: A Qualitative Analysis. Park. Dis. 2023, e5932865 (2023).

59. Cabrera, L. Y., Gilbert, M. M. C., McCright, A. M., Achtyes, E. D. & Bluhm, R. Beyond the Cuckoo’s Nest: Patient and Public Attitudes about Psychiatric Electroceutical Interventions. Psychiatr. Q. 92, 1425–1438 (2021).

60. Dalton, B. et al. ‘My dad was like “it’s your brain, what are you doing?”’: Participant experiences of repetitive transcranial magnetic stimulation treatment in severe enduring anorexia nervosa. Eur. Eat. Disord. Rev. 30, 237–249 (2022).

61. de Haan, S., Rietveld, E., Stokhof, M. & Denys, D. The phenomenology of deep brain stimulation-induced changes in OCD: an enactive affordance-based model. Front. Hum. Neurosci. 7, (2013).

62. Haan, S. de, Rietveld, E., Stokhof, M. & Denys, D. Effects of Deep Brain Stimulation on the Lived Experience of Obsessive-Compulsive Disorder Patients: In-Depth Interviews with 18 Patients. PLOS ONE 10, e0135524 (2015).

63. Leykin, Y. et al. Participants’ Perceptions of Deep Brain Stimulation Research for Treatment-Resistant Depression: Risks, Benefits, and Therapeutic Misconception. AJOB Prim. Res. 2, 33–41 (2011).

64. Sankar, T. et al. Deep Brain Stimulation Influences Brain Structure in Alzheimer’s Disease. Brain Stimulat. 8, 645–654 (2015).

65. Laxton, A. W. & Lozano, A. M. Deep Brain Stimulation for the Treatment of Alzheimer Disease and Dementias. World Neurosurg. 80, S28.e1–S28.e8 (2013).

66. Lam, J., Lee, J., Liu, C. Y., Lozano, A. M. & Lee, D. J. Deep Brain Stimulation for Alzheimer’s Disease: Tackling Circuit Dysfunction. Neuromodulation Technol. Neural Interface 24, 171–186 (2021).

67. Sattler, S. & Pietralla, D. Public attitudes towards neurotechnology: Findings from two experiments concerning Brain Stimulation Devices (BSDs) and Brain-Computer Interfaces (BCIs). PLOS ONE 17, e0275454 (2022).

68. MacDuffie, K. E., Ransom, S. & Klein, E. Neuroethics Inside and Out: A Comparative Survey of Neural Device Industry Representatives and the General Public on Ethical Issues and Principles in Neurotechnology. AJOB Neurosci. 13, 44–54 (2022).

69. Cabrera, L. Y., Achtyes, E. D., Bluhm, R. & McCright, A. M. Views about neuromodulation interventions for depression by stakeholder group, treatment modality, and depression severity. Compr. Psychiatry 122, 152365 (2023).

70. Bluhm, R., Sipahi, E. D., Achtyes, E. D., McCright, A. M. & Cabrera, L. Y. Stakeholders’ Ethical Concerns Regarding Psychiatric Electroceutical Interventions: Results from a US Nationwide Survey. AJOB Empir. Bioeth. 15, 11–21 (2024).

71. Lozano, A. M. et al. Deep brain stimulation: current challenges and future directions. Nat. Rev. Neurol. 15, 148–160 (2019).

72. Reddy, A. et al. Deep brain stimulation, lesioning, focused ultrasound: update on utility. AIMS Neurosci. 10, 87–108 (2023).

73. Merner, A. R. & Kubu, C. S. The Potential Harms of Speculative Neuroethics Research. AJOB Neurosci. 14, 418–421 (2023).

74. Brody, D. J. & Gu, Q. Antidepressant Use Among Adults: United States, 2015–2018. https://www.cdc.gov/nchs/data/databriefs/db377-H.pdf (2020).

75. Morris, M. R. et al. Use of psychiatric medication by college students: A decade of data. Pharmacother. J. Hum. Pharmacol. Drug Ther. 41, 350–358 (2021).

76. Bushnell, G. A. et al. Treating pediatric anxiety: Initial use of SSRIs and other anti-anxiety prescription medications. J. Clin. Psychiatry 79, 16m11415 (2018).

77. Bluhm, R., Cortright, M., Achtyes, E. D. & Cabrera, L. Y. “They Are Invasive in Different Ways.”: Stakeholders’ Perceptions of the Invasiveness of Psychiatric Electroceutical Interventions. AJOB Neurosci. 14, 1–12 (2023).

78. Wilkinson, S. T. et al. Barriers to the Implementation of Electroconvulsive Therapy (ECT): Results From a Nationwide Survey of ECT Practitioners. Psychiatr. Serv. 72, 752–757 (2021).

79. Chen, T. et al. Cost of Deep Brain Stimulation Infection Resulting in Explantation. Stereotact. Funct. Neurosurg. 95, 117–124 (2017).

80. Ooms, P. et al. Cost-effectiveness of deep brain stimulation versus treatment as usual for obsessive-compulsive disorder. Brain Stimulat. 10, 836–842 (2017).

81. Jameel, A. et al. The cost-effectiveness of unilateral magnetic resonance-guided focused ultrasound in comparison with unilateral deep brain stimulation for the treatment of medically refractory essential tremor in England. Br. J. Radiol. 95, 20220137 (2022).

